# On the mathematical modelling and analysis of Listeriosis from contaminated food products

**DOI:** 10.1101/2021.07.25.21261099

**Authors:** V. M. Mbalilo, C. W. Chukwu, F. Nyabadza

## Abstract

Listeriosis is a food-borne disease caused by a bacterium known as *Listeria monocytogens*. Humans can be infected by consuming contaminated food products. A transmission can also occur through contact with infected animals or people, however to a less extent. In this paper, a mathematical model for Listeriosis dynamics was developed. The steady states and their stability of the model system were determined and analyzed. The result shows that the disease-free equilibrium is asymptotically stable if the bacteria’s growth rate is less than its removal rate, and also the growth rate of food contamination is less than its removal rate. It was further observed that we can still have Listeriosis driven by the contaminated food products even if the Listeria bacteria population in the environment is very small.The results indicate that Listeriosis can be effectively controlled by removing contaminated food products, which was the policy adopted by the South African government during the recent Listeriosis outbreak.

## 1 Introduction

*Listeria monocytogens* are pathogens that are found in both raw and processed food products. The consumption of such products causes infections among humans and animals. This pathogen can be characterized as a Gram-positive bacteria [3]. Listeria strives very well in low-temperature areas and unclean environments. Other favourable conditions for their survival include high salt concentration and acidic conditions. Specifically, they are mostly found in soil, unclean water bodies (lakes, rivers, etc.), vegetation, refrigerators, faeces of some animals as well as in foods such as smoked fish, cold meats and soft cheeses. Animals such as cattle and poultry can also carry this bacteria. Due to their presence in soil and vegetation, they easily affect raw foods. Moreover, processed food can be contaminated by the presence of infected raw food and the use of unclean processing materials [15]. Humans can be infected by consuming such contaminated food products. However a transmission can also occur through contact with infected animals or people although to a less extent. Infected pregnant women and female animals can also transfer these infections to their unborn babies [22], a process often referred to as vertical transmission.

The group of people who are at high risk to acquire this disease are elderly, pregnant women and infants, as well as people with weak immune systems caused by illnesses with illnesses such as cancer, diabetes, kidney disease and HIV AIDS patients [7]. Fever, flu-like symptoms, vomiting, nausea and diarrhoea are the main symptoms of Listeriosis. Like most bacterial infections, Listeriosis can be treated and prevented by storing food safely, avoiding storing products in the fridge beyond the use-by date, cooking the meat and poultry properly and keeping raw food from touching other foods and utensils [19]. Although the disease is relatively rare, it is often severe with high hospitalisation and mortality rates. For instance, there were 10 cases of Listeriosis reported from a small area of Switzerland which was due to the distribution of local soft cheese. The same problem was experienced by the Czech Republic in 2006, with 78 patients of whom 13 died. Also, in 2006 and 2007, Germany reported having an outbreak of 16 cases caused by pre-sliced ready-to-eat meat products [16]. Furthermore, the National Institute of Communicable Disease (NICD) in South Africa reported that by 28 February 2018 there had been 943 laboratory-confirmed cases of Listeriosis with 176 deaths from the disease [4].

Mathematical models have been used to analyse the transmission dynamics of infectious diseases in the past. Several mathematical models and statistical methods have been used to study the spread of Listeriosis, see for instance [9–13]. The work by Buchanan *et al*. [8] shows that the growth of *Listeria monocytogens* depends on the interaction of five variables namely pH, temperature, sodium nitrate, atmosphere, and sodium chloride. According to their data, they found out that sodium nitrate can have high significant bacteriostatic activity against *Listeria monocytogens* and hence can be used to provide cured meats with a degree of protection against this bacteria. This is achieved when there is a combination of high salt concentration, acidic pH, vacuum packaging and adequate refrigeration. Ivanek *et al*. [5] conducted a study on how ready-to-eat food (smoked fish) is contaminated with *Listeria monocytogens* in relation to some associated factors during food processing. In their study, they developed a model where they considered food contact surfaces and employees gloves as key factors for food contamination. It was discovered that, the best way to prevent food contamination during food processing is to make sure that the raw food and processing materials coming in are free from contamination. According to the inference from the outbreak of the disease reported in Japan, Europe and North America, the foods which are at high risk for susceptible humans are ready-to-eat meats and soft cheese [20]. Furthermore, Luber *et al*. [14] pointed out that for successful employment of the food safety methods against Listeriosis there is a need for food workers to be educated and trained on how to prevent (protect) the ready-to-eat foods from Listeria monocytogens contamination and also to advise the consumer to be aware and responsible for their food safety. In [17], conducted a study with the aim of developing a predictive model that simulates the growth of *Listeria monocytogene* in soft blue-white cheese. In their study, they come up with a tertiary predictive model of the Listeria growth as a function of lactic acid, temperature, pH and sodium chloride. According to their analysis, they found out that the growth rate of *Listeria monocytogenes* is very high when present in cheese. Mateus *et al*. [1] pointed out that pregnant women affected with Listeriosis are at a high risk of miscarriages and fetal death or neonatal morbidity in the form of meningitis and septicemia. They also found out that improving education about Listeriosis transmission, control and prevention for pregnant and individuals with weak immune systems will help in minimizing the mortality rate of this disease. Moreover, [13] developed a mathematical model to study the effect of the vaccination of animals on the spread of Listeriosis among humans and animals (as vectors of Listeria). In their analysis, they found out that, secondary infections can increase due to the decrease of human and animal death rate as well as animal recovery rate.

Very few mathematical models the role of the environment and contaminated food products have been developed to date. Simple model that compute the role of bacteria and food products in transmission of dynamics of Listeriosis in the human population is formulated with the aim of investigating the role of the few control measures that are available in the event of the out break. The control measure include reduction of the rate of infection, removal of contaminated food product and hygiene in the food manufacturing process. While the model presented in this page represent the simplest caricature of the infection dynamics of Listeriosis, the results have significant influence on the management and qualification of control strategies over the long period of time. We ague that despite its simplest, the model offers significant insights in the modelling of Listeriosis driven by contaminated food products and bacteria from the environment.

The outline of this paper is as follows; Section 1 introduces the research paper followed by a model formulation in Section 2. The model basic properties and analyses are presented in Section 3. Numerical simulations are presented in Section 4 and Section 5 concludes the paper.

## 2 Model Formulation

In this section, we develop a mathematical model for Listeriosis dynamics which is divided into three components namely; the human population, Listeria and factory products. The human population comprises of three compartments which are susceptible humans, *S*(*t*), infected humans, *I*(*t*), and the recovered humans, *R*(*t*). Individuals are recruited into susceptible class at a rate proportional to the total human population *N* (*t*) so that the recruitment is modelled by of *µN*, where *µ* is the natural birth/mortality rate. The susceptible humans can move into infected class either by acquiring Listeriosis through eating contaminated products or through contact with contaminated material from the environment with a force of infection *λ*_*h*_(*t*), defined after system (2). Furthermore, the infected humans can either die naturally, die of the disease at a rate of *δ*_1_ or recover at a rate of *γ* and join the recovered class. We are assuming that there is no human to human transmission. Considering the factory dynamics, we are assuming that the amount of food products, *F* (*t*) comprises of non-contaminated, *F*_*n*_(*t*), and contaminated, *F*_*c*_(*t*) food products. We assume that factory products are manufactured or produced at a rate of *δ*_2_*F*. By contact with contaminated surfaces and contaminated products, the non-contaminated products can become contaminated with Listeria bacteria. Non-contaminated factory products become contaminated with a force of infection *λ*_*f*_ (*t*) defined after system (2). The Listeria bacteria in the environment is assumed to grow at a rate of *r*_*b*_, and die at a rate *µ*_*b*_. The growth of the bacteria is assumed to be logistic with a carrying capacity of *K*. We assume that, at any time, *t*, the human population and factory food products are constant and respectively given by;

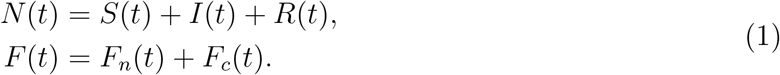

The model structure is represented schematically in Figure 1. From the above model description and Figure 1 we obtain the following system of non-linear ordinary differential equations:

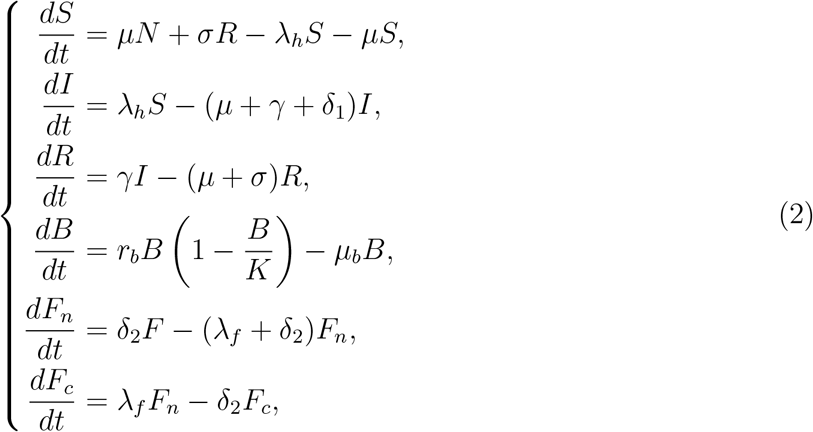

where

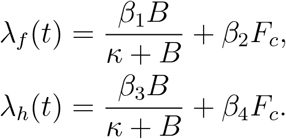

We rescale system (2) by setting

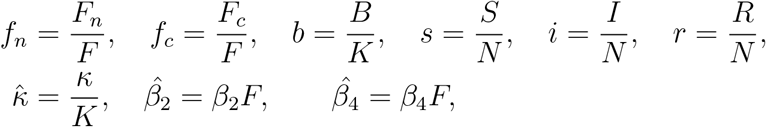

and substituting *r* = *n* − *s* − *i*, to obtain the following system of differential equations:

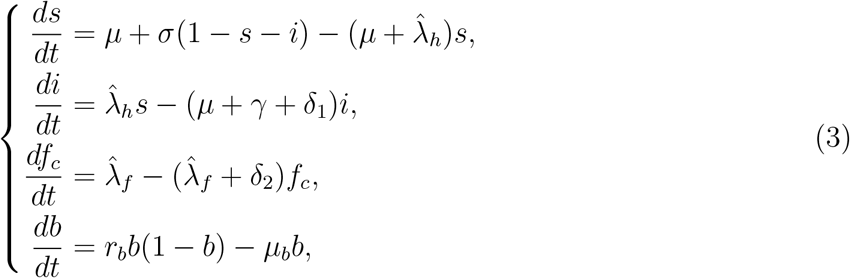

where re-scaled forces of infections are

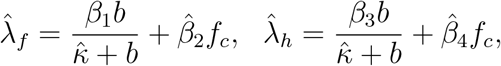

subject to the following initial conditions

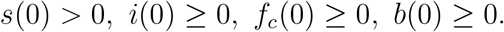

**Figure 1:**
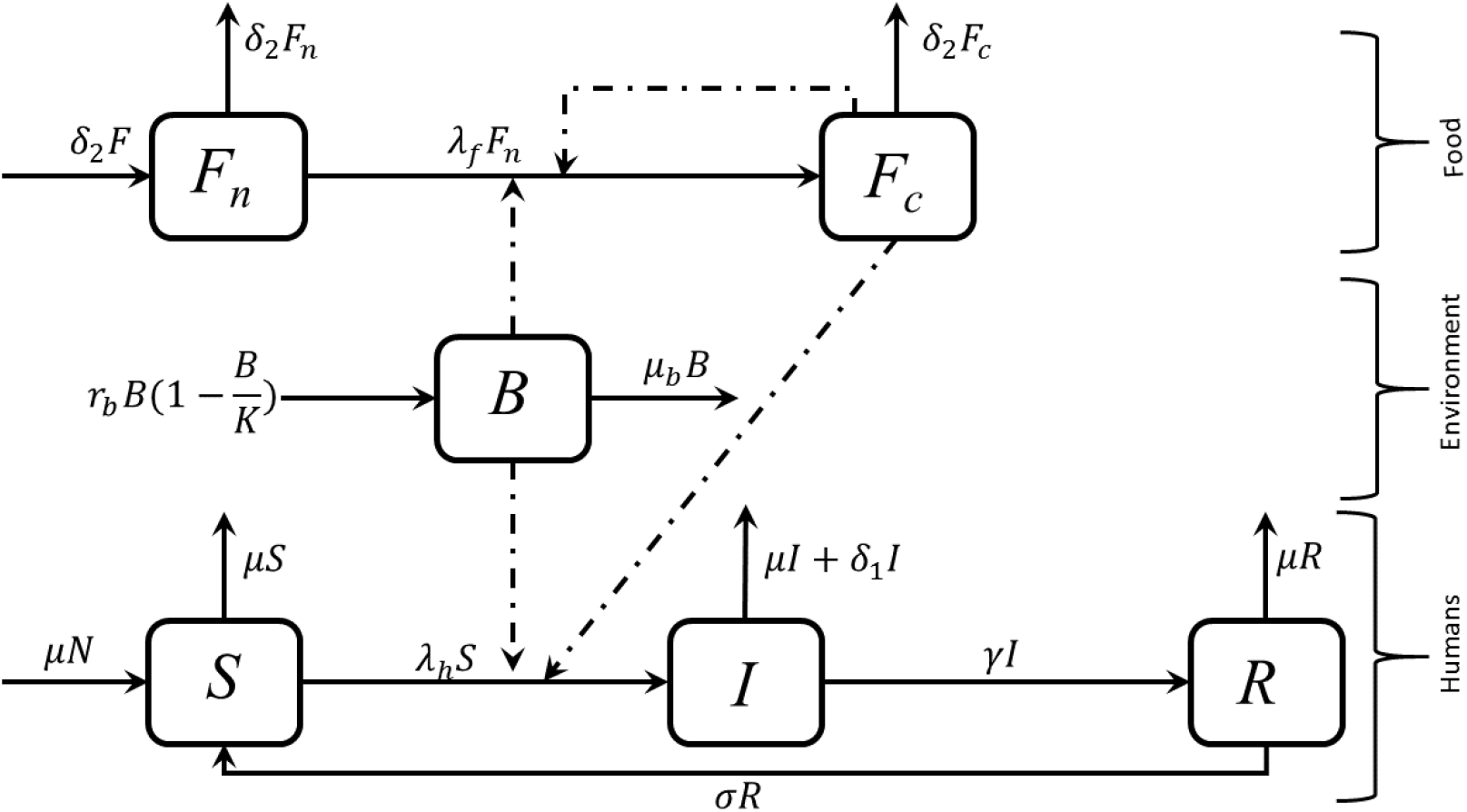
Model flow chart for Listeriosis disease transmission dynamics. The dotted and solid line shows the contributing factors for the human and food products to become contaminated as well as infection links respectively.

All parameters of model system (3) are assumed to be positive at all time *t >* 0.

### 2.1 Basic Properties

We show that all the solutions of model system (3) has non-negative solutions, and bounded for *t >* 0, that is remain biologically meaningful into feasible region (Ω).

#### 2.1.1 Positivity of Solutions

The positivity of solutions to our model is given by the following theorem.

##### Theorem 1.

*Suppose s*(0) *>* 0, *i*(0) ≥ 0, *f*_*c*_(0) ≥ 0, *b*(0) ≥ 0 *then the solutions of s*(*t*), *i*(*t*), *f*_*c*_(*t*) *and b*(*t*) *of system (3) are non-negative for all time t* ≥ 0.

*Proof*. Let (*s*(*t*), *i*(*t*), *f*_*c*_(*t*), *b*(*t*)) be a solution of the system (3) with the given initial conditions. From the first equation of system (3), we have that

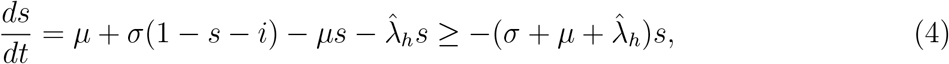

is the differential inequality from (4). Integration yields

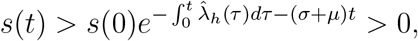

where *s*(0) is the given initial condition for *s*(*t*). From the second equation of model equation (3) we have the differential inequality

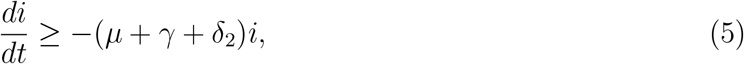

so that

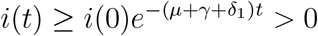

where *i*(0) is the initial condition for *i*(*t*). The remaining equations yields

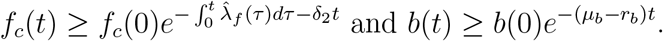

Thus, the solutions (*s*(*t*), *i*(*t*), *f*_*c*_(*t*), *b*(*t*)) will remain positive in Ω at all time *t* ≥ 0.

#### 2.1.2 Feasible Region

The feasible region of our model is captured by the following theorem.

##### Theorem 2.

*Consider the biologically feasible region given by*

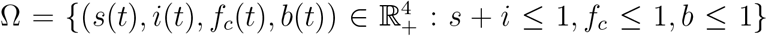. *The solution of the system (3) with the non-negative initial condition are bounded for all t ≥* 0 *in the biologically feasible region*, (Ω).

*Proof*. The total change in the human population is given by

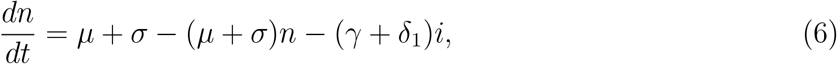

where *n* = (*s* + *i*) ≤ 1. However in the absence of mortality due to human Listeriosis infections, equation (6) reduces to

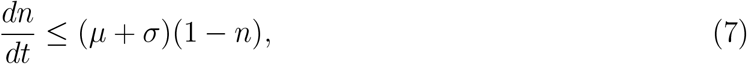

whose solutions yields

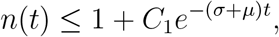

where *C*_1_ is a constant. We note that as *t* → ∞, then *n*(*t*) → 1.

Considering the contaminated food product given by last equation of system (3), we have that

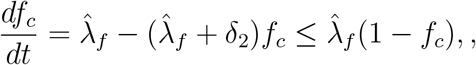

whose solution yields

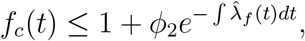

where *ϕ*_2_ is a constant. Note that as *t* → ∞ we have *f*_*c*_(*t*) → 1.

Furthermore, for the bacteria population, considering the third equation of system (3), we have the Bernoulli’s equation

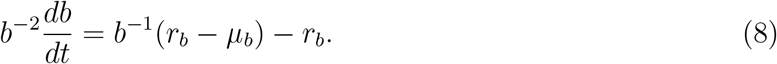

The solution of equation (8) we obtain

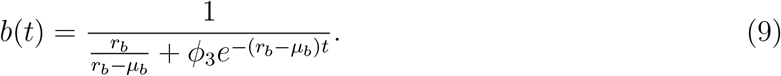

Thus, as *t* → ∞, 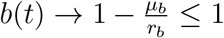. So, we note that for the Listeria to exist its death rate (*µ*_*b*_) must be less than the growth rate (*r*_*b*_), which implies that, 0 ≤ *b* ≤ 1.

Hence (*s, i, f*_*c*_, *b*) are all bounded in region Ω, and are biologically feasible, which complete the proof.

### 2.2 Model Steady States

We determine the steady states of system (3) by setting the right hand side to zero as follows

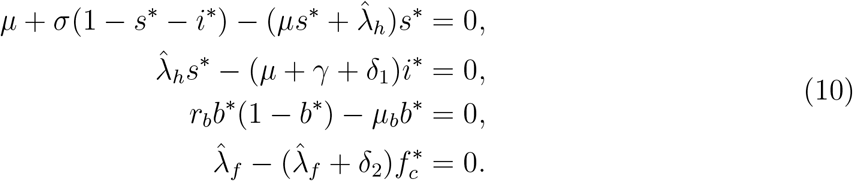

From the third equation of the system (10), we have that

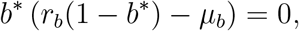

which gives

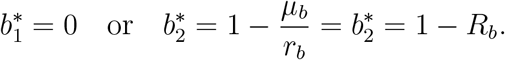

where

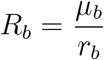

#### Remark 1.

*Note that*, 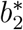 *exists if and only if R*_*b*_ *<* 1.

We consider two cases.

**CASE 1:** When 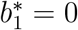, we have that

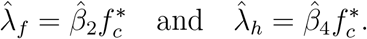

Now from the last equation of the system (10) we have

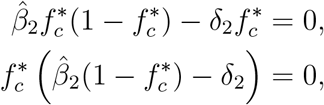

and this yields

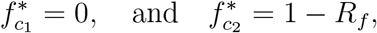

where

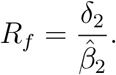

#### Lemma 1.

*The steady state* 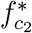 *exist if and only if R*_*f*_ *<* 1.

Now if *b*^*^ = 0, then 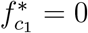, which implies that from the second equation of (10). Now substituting into first equation of the system (10) we have

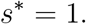

Therefore, we have a disease free state (DFS) denoted by

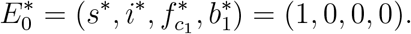

From the second equation of the system (10) we have

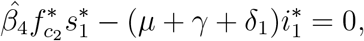

which yields

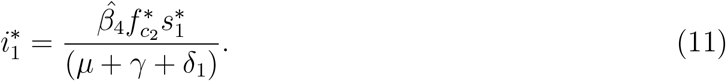

Substituting equation (11) into the first equation of the system (10) we have

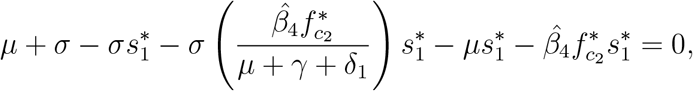

which yields

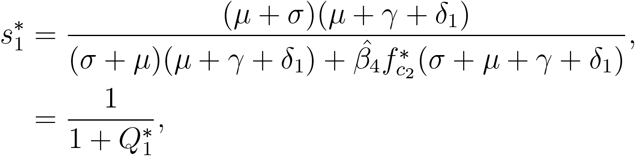

after some algebraic manipulation where 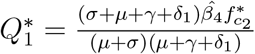.

Substituting 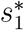 into (11) we have that

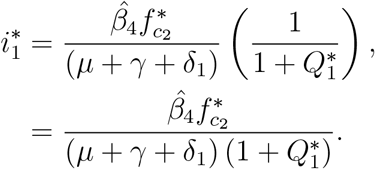

This results in the bacteria free state (BFS)

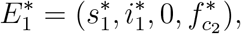

**Case 2:** If 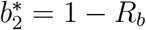; then,

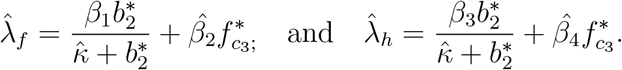

Similarly from the last equation of the system (10), we have that

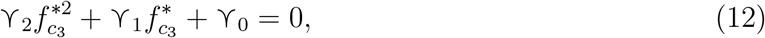

after some algebraic simplification, where

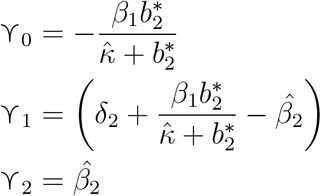

Solving equation (12) we have,

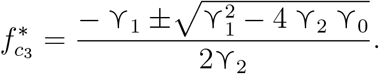

Since ϒ _0_ *<* 0 and ϒ _2_ *>* 0 then ϒ _2_ ϒ _0_ *<* 0, and this implies that 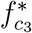 has one positive solution, say 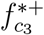 irrespective of the conditions imposed on ϒ _1_.

Now from the second equation of the system (10), we have

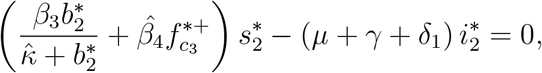

which yields

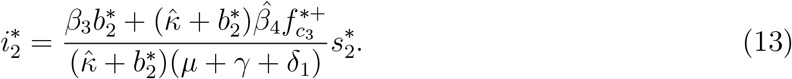

Further, from the first equation of the system (10), we have

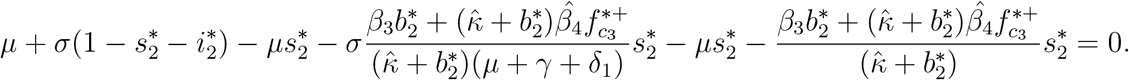

This give

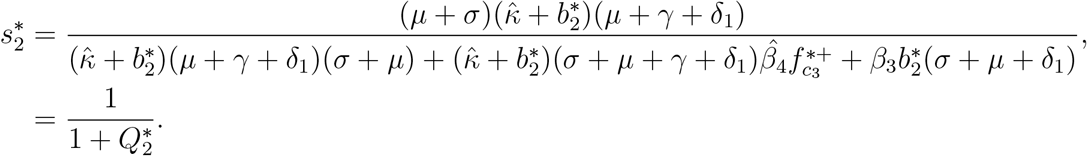

where

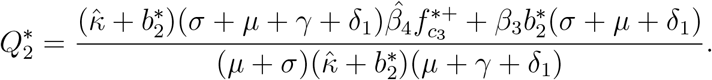

Next, we substitute 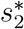 into equation (13) and obtain

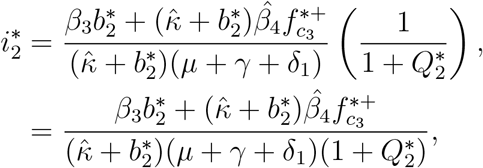

This results to endemic state (ES) given by

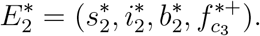

## 3 Stability Analyses

In this section we prove the local stability of the steady states by using the Jaccobian matrix linearized at each steady state.

### 3.1 Local Stability of the DFS

#### Theorem 3.

*The disease free state* 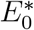, *is locally asymptotically stable if R*_*f*_ *>* 1 *and r*_*b*_ *< µ*_*b*_.

*Proof*. In order to determine the local stability of the disease free steady state we evaluate the Jaccobian matrix at 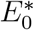 which gives

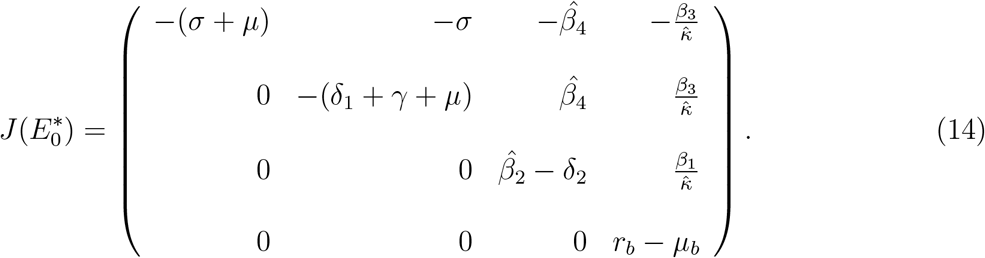

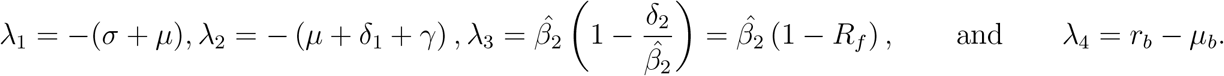

Since all parameters are positive, it is clear that *λ*_1_ and *λ*_2_ are negative. However, *λ*_3_ and *λ*_4_ are negative if and only if *R*_*f*_ *>* 1 and *r*_*b*_ *< µ*_*b*_ respectively. Therefore, 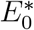, is locally asymptotically stable if *R*_*f*_ *>* 1 and *r*_*b*_ *< µ*_*b*_.

**Remark 2.** *The growth rate of food contamination must be less than its removal and the growth rate of bacteria must be less than its removal (natural death rate of bacteria).*

### 3.2 Local Stability of the BFS

#### Theorem 4.

*The bacteria free state* 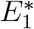 *is locally asymptotically stable if r*_*b*_ *< µ*_*b*_ *and R*_*f*_ *<* 1.

*Proof*. We evaluate the Jacobian matrix at 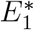 and obtain

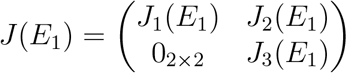

where

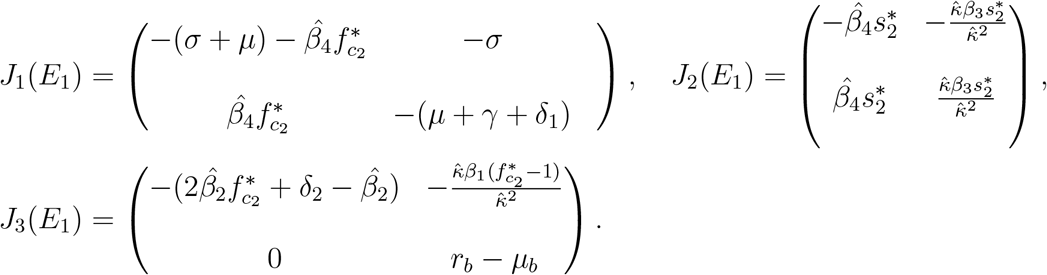

The eigenvalues from *J*_3_(*E*_1_) are;

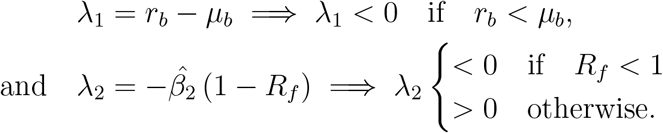

Further, the eigenvalues from *J*_1_(*E*_1_) is given by the characteristic polynomial

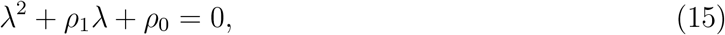

where

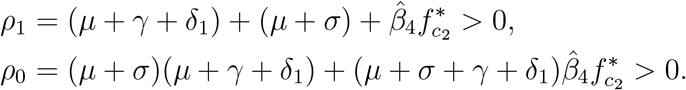

Since *ρ*_1_ *>* 0 and *ρ*_0_ *>* 0, the eigenvalues of equation (15) have negative real parts by the Routh–Hurwitz stability criterion.

Therefore, the bacteria free state is locally asymptotically stable if and only if *µ*_*b*_ *> r*_*b*_ and *R*_*f*_ *<* 1.

### 3.3 Local Stability of the ES

#### Theorem 5.

*The endemic state* 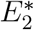 *is locally asymptotically stable if* 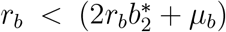 *and* 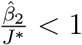, *where*

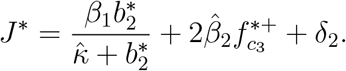

*Proof*. We evaluate the Jacobian matrix at 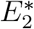 to obtain

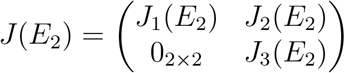

where

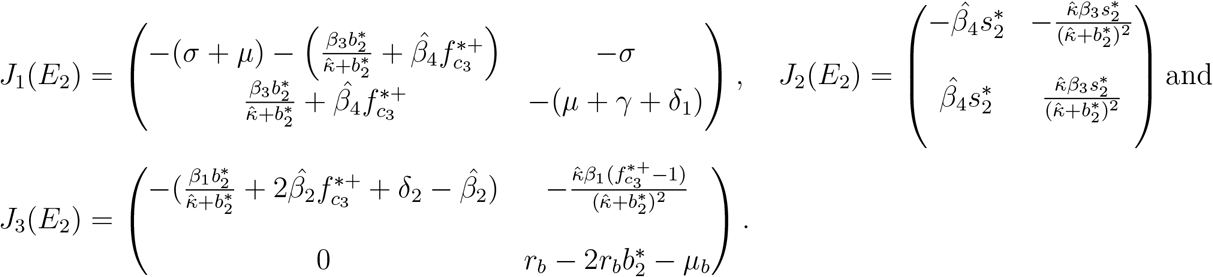

The eigenvalues from *J*_3_(*E*_2_) are;

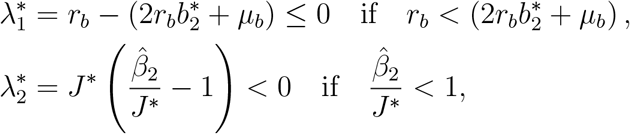

where

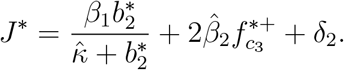

Further, the eigenvalues of matrix *J*_1_(*E*_2_) is given by the characteristic polynomial

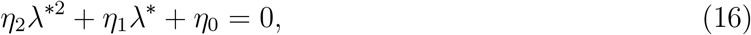

where

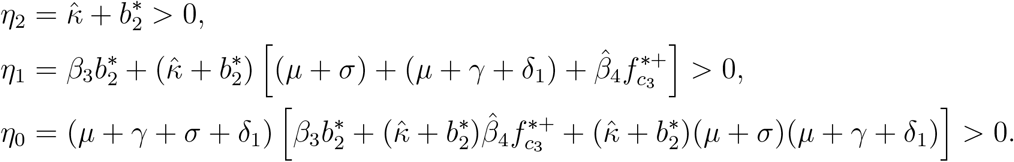

So, equation (16) can be rewritten as

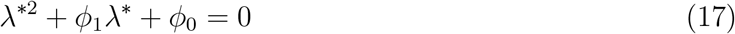

where 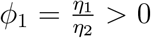 and 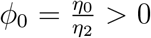. Since *ϕ*_1_ *>* 0 and *ϕ*_0_ *>* 0, The eigenvalues of equation (17) have negative real parts by the Routh-Hurwitz stability criterion. Therefore, the endemic state is locally asymptotically stable if 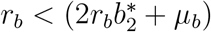 and 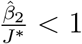.

## 4 Numerical Simulations

### 4.1 Parameter Values

In this section, we simulate our model for Listeriosis in order to determine the time series trajectories of the disease in human population. The system of equations were solved by using Julia and Matlab software over a period of time using the estimated parameter values in Table 2. We hypothetically chose the parameter values in such a way that they yield the results of the analyses of the steady states. The parameter values in Table 2 are all assumed since there are very few mathematical models on human Listeriosis disease infection and in literature. A summary descriptions of the parameters used in the model are shown in Table 2 at the endemic state with initial conditions *s*(0) = 0.4, *i*(0) = 0.1, *f*_*c*_(0) = 0.2, *b*(0) = 0.0005.

**Table 1:** Parameter values for simulations.

| Parameter | Symbols | Values(day<sup>-1</sup>) | Ref. |
| --- | --- | --- | --- |
| Human natural death rate | $\mu$ | 0.02 | Estimated |
| Human recovery rate | $\gamma$ | 0.07 | Estimated |
| Rate loss of immunity | $\sigma$ | 0.99 | Estimated |
| Growth rate of Listeria | $r_b$ | 0.32 | [12] |
| Natural death rate of bacteria | $\mu_b$ | 0.2-0.3[0.2] | [25] |
| Disease induced death rate | $\delta_1$ | 0.004 | Estimated |
| Rate of removal of food products | $\delta_2$ | 0.017 | |
| Half saturation constant | $\hat{\kappa}$ | 0.0002 | Estimated |
| Rate at which non-contaminated food are contaminated by bacteria | $\beta_1$ | 0.004 | Estimated |
| Rate of contamination of food products | $\hat{\beta}_2$ | 0.006 | Estimated |
| Infection rate of humans by bacteria from the environment | $\beta_3$ | 0.03 | Estimated |
| Infection rate of susceptible humans by contaminated food products | $\hat{\beta}_4$ | 0.05 | Estimated |

**Table 2:** PRCC and P-values.

| Parameter | P-values | PRCC values |
| --- | --- | --- |
| $\mu$ | 0.0230 | -0.1969 |
| $\gamma$ | 0.0788 | -0.2984 |
| $\sigma$ | 0.9536 | 0.0927 |
| $r_b$ | 0.1848 | 0.7602 |
| $\mu_b$ | 0.2157 | -0.6504 |
| $\delta_1$ | 0.0040 | -0.0504 |
| $\delta_2$ | 0.0183 | -0.0706 |
| $\hat{\kappa}$ | 0.0001 | -0.0798 |
| $\beta_1$ | 0.0042 | 0.1708 |
| $\hat{\beta}_2$ | 0.0067 | 0.0663 |
| $\beta_3$ | 0.0177 | 0.5641 |
| $\hat{\beta}_4$ | 0.0445 | 0.2388 |

### 4.2 Convergence of Steady State

Figure 2(a) depicts the graph of the equilibrium point 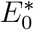 in which all the infected compartments tends to zero. Therefore this shows the stability of the disease free equilibrium point. We can infer that this is due to the high death rate, *µ*_*b*_ = 0.4, of the bacteria as compared to their growth rate *r*_*b*_ = 0.18. So there are less infections caused by the bacteria. Moreover, the rate of removal of contaminated food, *δ*_2_ = 0.5, is higher as compared to the growth rate of food contamination, 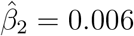. Figure 2(b) depicts the graph of the equilibrium point 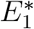. We can observe from this graph that the bacteria population goes to extinct since their growth rate is less that their death rate. However, Listeriosis infections exist in the population due to presence of contaminated food. This can be attributed to the fact that the rate of removal of contaminated food, *δ*_2_ = 0.1, is less as compared to the growth rate of food contamination, 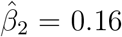. Figure 2(c) shows the graph of the endemic equilibrium point 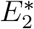 in which the contaminated products, the bacteria in the environment and the human population are all non-zero. Hence we can observe that the disease will persist.

**Figure 2:**
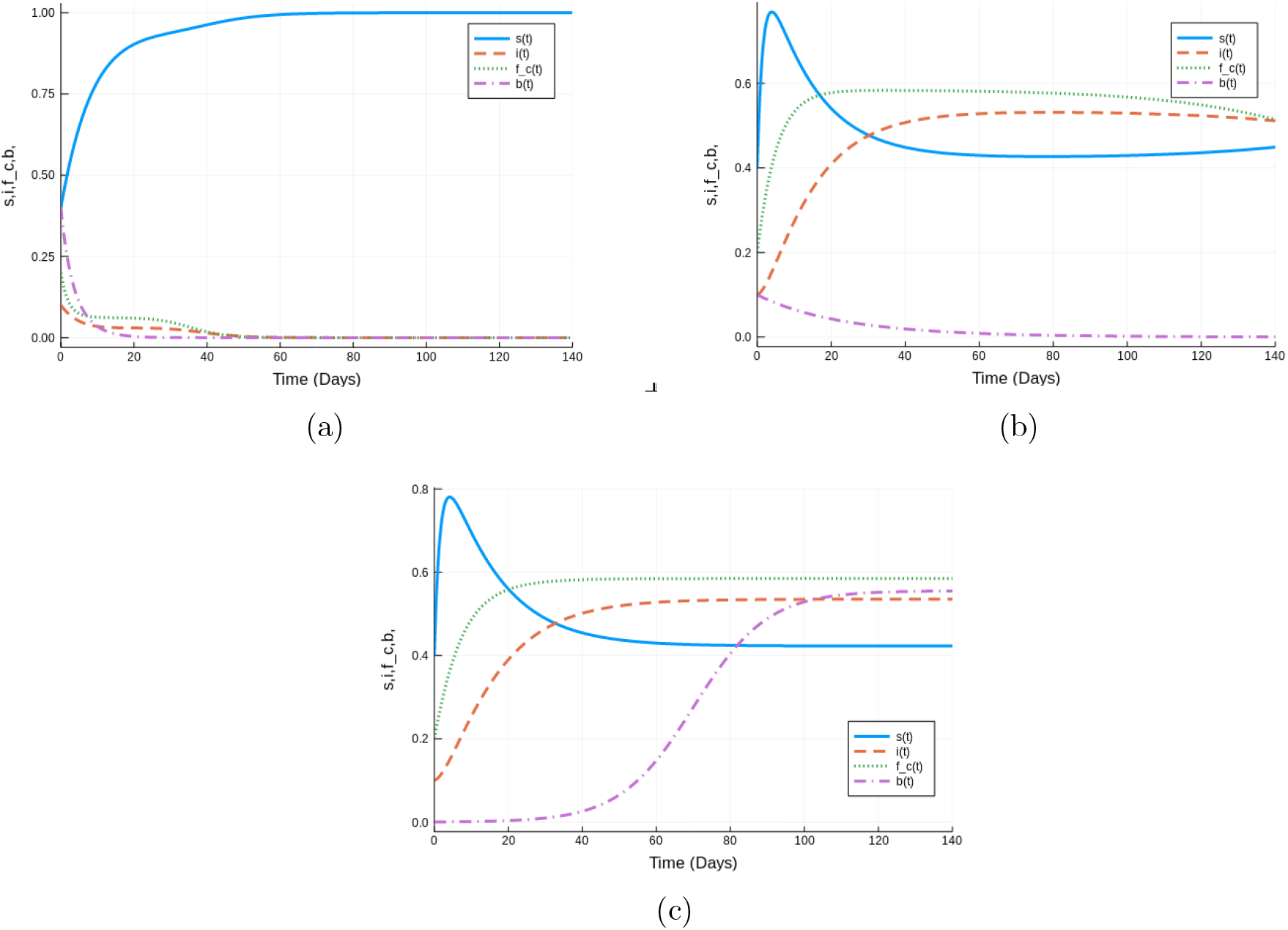
(a) The steady state 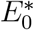 with parameter values *µ* = 0.028, *σ* = 0.1, *γ* = 0.08, *δ*_1_ = 0.1, *δ*_2_ = 0.5, *r*_*b*_ = 0.18, *µ*_*b*_ = 0.4, *β*_1_ = 0.004, 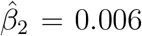, *β*_3_ = 0.03, 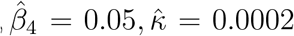 and initial conditions *s*(0) = 0.4, *i*(0) = 0.1, *f*_*c*_(0) = 0.2, *b*(0) = 0.4. (b) The steady state 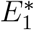 with parameter values, *µ* = 0.028, *σ* = 0.6, *γ* = 0.004, *δ*_1_ = 0.0035, *δ*_2_ = 0.1, *r*_*b*_ = 0.05, *µ*_*b*_ = 0.089, *β*_1_ = 0.004, 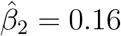, *β*_3_ = 0.1, 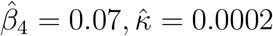 and initial conditions *s*(0) = 0.4, *i*(0) = 0.1, *f*_*c*_(0) = 0.2, *b*(0) = 0.1. (c) The steady state 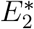 with parameter values in Table 2

### 4.3 Sensitivity Analysis

Global sensitivity analysis is a statistical tool used to determine the sensitivity and uncertainly of epidemiological parameters. Here, we employ the method of Latin-hyper cube sampling as in [24] to determine model parameters driving Listeriosis subject to the model under investigation. According to [24], for any modelling exercise, we can carry out sensitivity at a time point of interest or over a range of time frames to determine the sensitivity of the model parameters. Since humans can devolve fully Listeriosis infections between 1 to 90 days after consuming contaminated food products contaminated by Listeria. We carry out the simulation implemented in Matlab with a time step of 1 and between 1 to 90 days, [25], with a time average of 45 days as it is assumed to be an average time an infected person with Listeria should have to develop the symptom of the disease. The simulation result gives the partial Rank Correlation Coefficient’s (PRCCC) and P-values shown in Table 2 with the Tornado plot showing different parameters sensitivity values as shown in Figure 3. We observe in Figure 3, that, the parameters 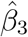 and *r*_*b*_ are strongly positively correlated while *µ*_*b*_ is strongly negatively correlated.. Hence, the increase in the food contamination rate 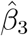 and the bacteria growth *r*_*b*_ will impact the disease variability leading to more infected humans in the community affected by Listeriosis. An increase in the removal rate of bacteria, *µ*_*b*_, will certainly result in fewer human infections.

**Figure 3:**
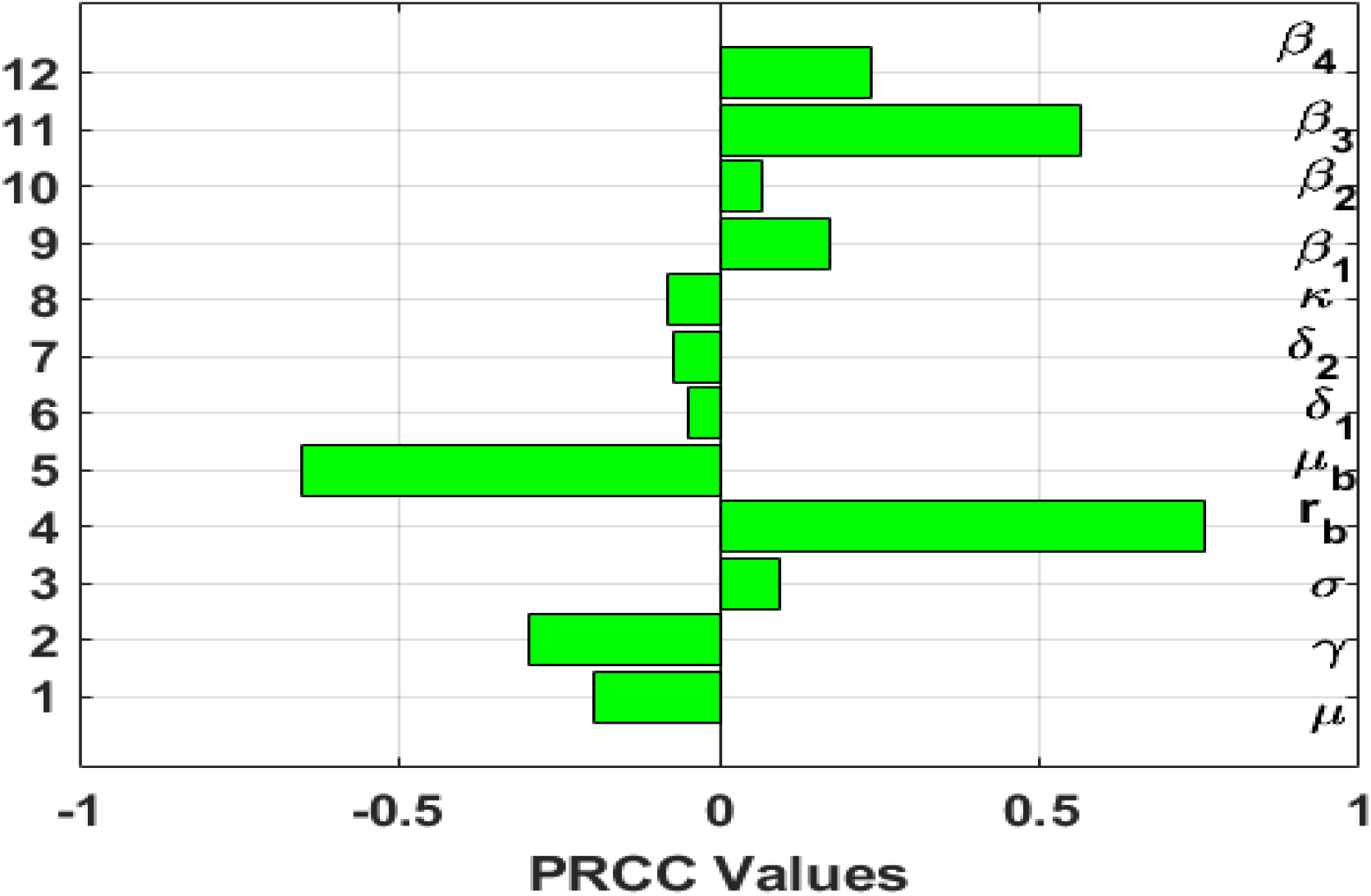
Tonardo plot showing different parameter PRCC values. Note that 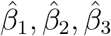 and 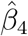 are represented by *β*_1_, *β*_2_, *β*_3_ and *β*_4_ respectively as Matlab do not compile the^command.

### 4.4 Effects of *δ*_2_ and *β*_2_ on Contamination Threshold *R*_*f*_

Figure 4(a) depicts the behaviour of contamination threshold *R*_*f*_ against the rate of contamination of food products, 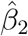, the graph shows that as the rate of contamination of food products, 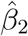, increase then the contamination threshold *R*_*f*_ decrease. Figure 4(b) depicts the behaviour of contamination threshold, *R*_*f*_, against the disposal rate *δ*_2_, the graph shows that as disposing rate *δ*_2_ increase then the contamination threshold, *R*_*f*_, also increase.

**Figure 4:**
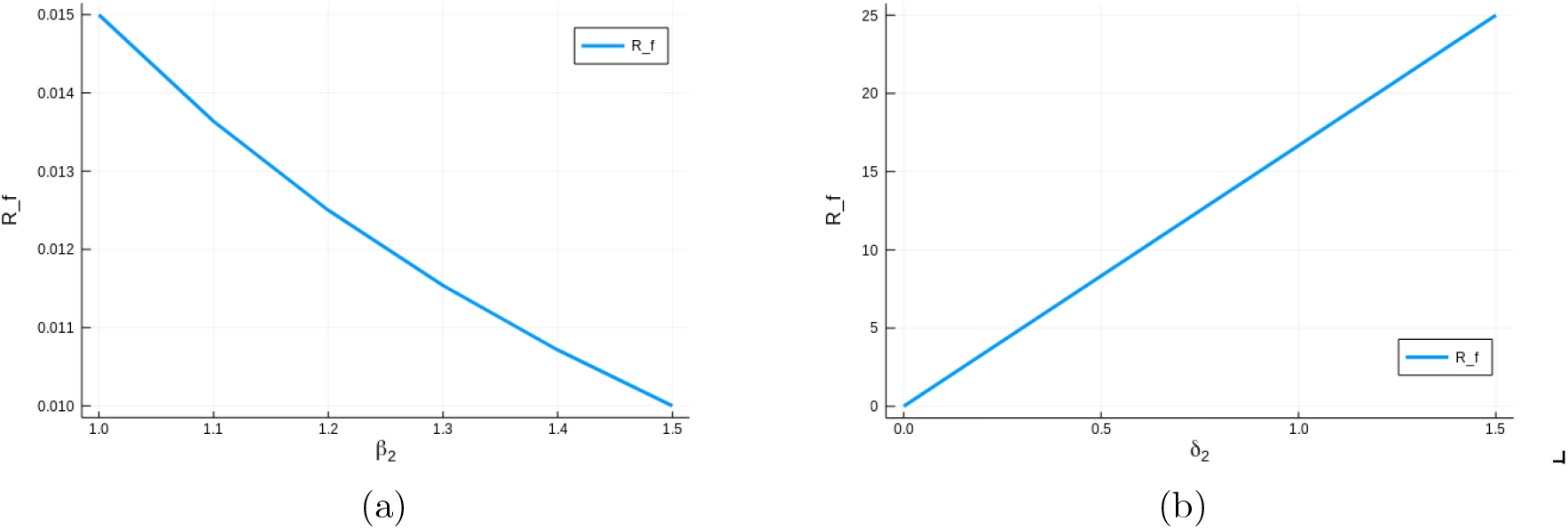
(a) The contamination threshold *R*_*f*_ with parameter values, *δ*_2_ = 0.015, 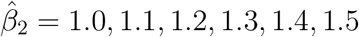. (b) The contamination threshold *R*_*f*_ with parameter values, 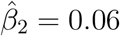, *δ*_2_ = 0.0, 1.0, 1.1, 1.2, 1.3, 1.4, 1.5.

### 4.5 Effects of Varying Parameters *δ*_2_, *β*_2_, *β*_3_ and *β*_4_ on Infected Humans

Figure 5(a) shows how infectious human decrease as disposing (removal) rate increase (*δ*_2_). Figure 5(b) shows how infectious human decrease as the bacterial natural death (removal) rate *µ*_*b*_ increases.

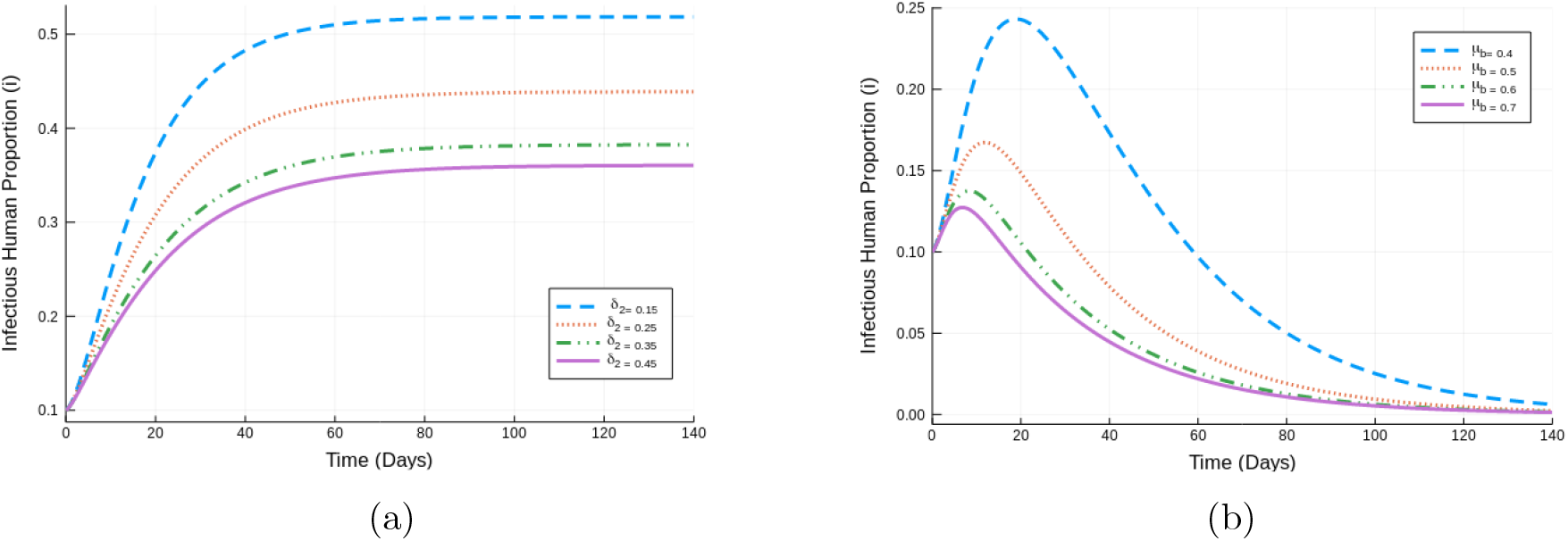

Figure 5(c) shows how infectious humans decrease as Listeria bacteria infection rate from the environment decrease. Figure 5(d) shows how infectious human decrease as the rate of food contamination *β*_4_, decreases.

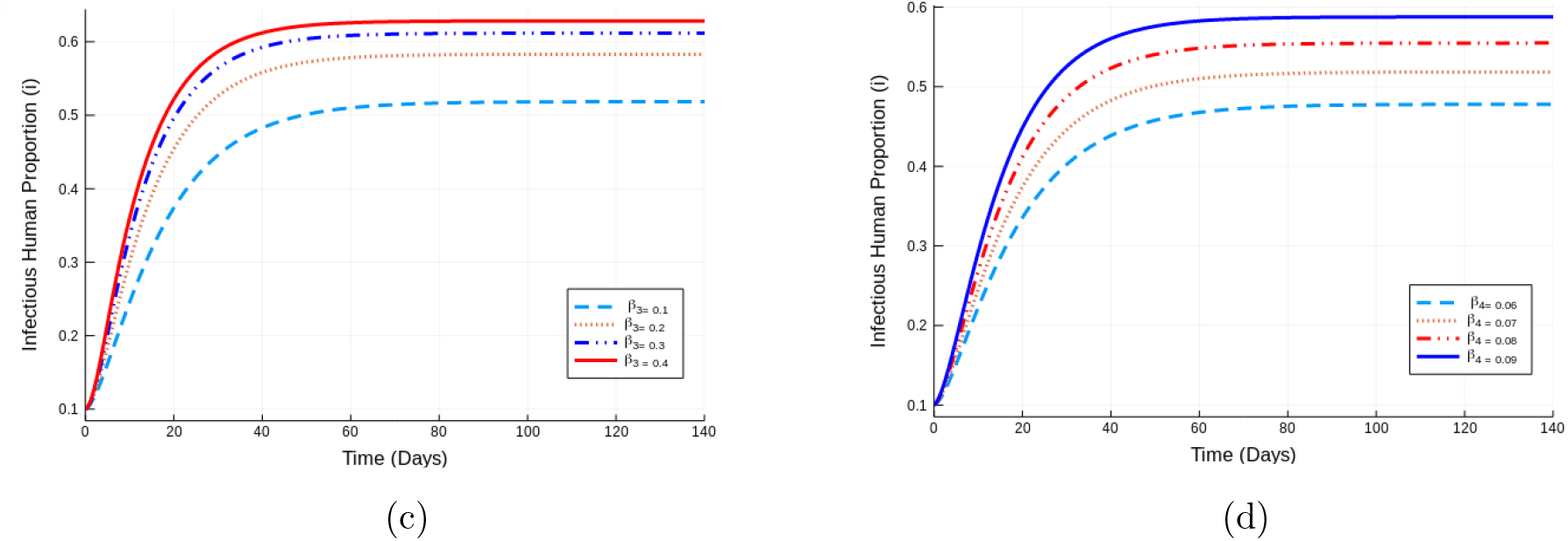

## 5 Conclusion

In this paper, we formulate a mathematical model to eludidate the contribution of contaminated food products and bacteria in the environment on the spread of Listeriosis. The model steady states were determined and their stabilities determined. The model has three steady states, the disease free steady states*E*_0_ in which their in only the susceptible population while the remaining state variables are zeros. While this is mathematically plausible this steady state may not be realistic as the bacteria will always be present in the environment. The second steady state is bacteria free steady state *E*_1_ in which the is driven by only contaminated food products. This steady state is driven by contamination within the food manufacturing environment. The third steady is the endemic state in which all the state variable are no-zero. This steady state depicts a scenario in which the environment, the human population and food products are active in spread of Listeriosis.

The model simulation are carried out to investigate the role of interventions in the spread of Listeriosis. The most important interventions in the removal of contaminated food product, modelled by the parameters *δ*_2_. Sensitivity analysis is also carried to determine the most significant parameters that influence the transmission dynamics. It is through these parameters that interventions can be designed and quantified.

## Data Availability

None

## Data Availability

No data used for this research work

## Acknowledgements

The authors would like to thank the Faculty of Science in the University of Johannesburg and AIMS South Africa for their support in completing this project.

